# Confirmed SARS-CoV-2 infection in Scottish neonates 2020-2022: a national, population-based cohort study

**DOI:** 10.1101/2022.07.28.22278152

**Authors:** Anna Goulding, Fiona McQuaid, Laura Lindsay, Utkarsh Agrawal, Bonnie Auyeung, Clara Calvert, Jade Carruthers, Cheryl Denny, Jack Donaghy, Sam Hillman, Lisa Hopcroft, Leanne Hopkins, Colin McCowan, Terry McLaughlin, Emily Moore, Lewis Richie, Colin R Simpson, Bob Taylor, Lynda Fenton, Louisa Pollock, Christopher Gale, Jennifer J Kurinczuk, Chris Robertson, Aziz Sheikh, Sarah Stock, Rachael Wood

## Abstract

**Objective:** To examine infants in Scotland aged 0-27 days with confirmed SARS-CoV-2 infection; the risk of neonatal infection by factors including maternal infection status and gestation at birth; and the need for hospital admission among infected neonates.

**Design:** Population-based cohort study.

**Setting and population:** All live births in Scotland, 1 March 2020 to 31 January 2022.

**Results:** There were 141 neonates with confirmed SARS-CoV-2 infection over the study period, giving an overall infection rate of 153 per 100,000 live births (141/92,009). Among infants born to women with confirmed infection around the time of birth, the infection rate was 1,811 per 100,000 live births (15/828). Nearly two-thirds (92/141, 65.2%) of babies with confirmed neonatal infection had an associated admission to neonatal or (more commonly) paediatric care. Of those admitted to hospital, 6/92 (6.5%) infants were admitted to neonatal or paediatric intensive care, however none of these six had COVID-19 recorded as the main diagnosis underlying their admission. There were no neonatal deaths among babies with confirmed infection.

**Implications and relevance:** Confirmed neonatal SARS-CoV-2 infection is uncommon. Secular trends in the neonatal infection rate broadly follow those seen in the general population, albeit at a lower level. Maternal infection at birth increases the risk of neonatal infection, but most babies with neonatal infection are born to women without confirmed infection. A high proportion of neonates with confirmed infection are admitted to hospital, with resulting implications for the baby, family, and services, although their outcomes are generally good.

**Key messages:** *What is already known on this topic:*

- The incidence of SARS-CoV-2 infection in neonates is low, but some studies have suggested that age under 1 month is a risk factor for severe infection requiring admission to intensive care.
- Almost all the studies of neonatal SARS-CoV-2 have focused on the transmission risk from SARS-CoV-2 positive women to their offspring and data are lacking on the level of neonatal SARS-CoV-2 infection in the whole population.

*What this study adds:*

- This study includes all babies with confirmed SARS-CoV-2 in the neonatal period in Scotland during the first 22 months of the COVID-19 pandemic.
- Confirmed neonatal SARS-CoV-2 infection is uncommon, but a high proportion of neonates with confirmed infection are admitted to hospital.
- Confirmed maternal SARS-CoV-2 infection around the time of birth substantially increases the risk of neonatal infection, although the absolute risk of neonatal infection remains low (<2%) and most babies with neonatal infection are born to women without confirmed infection.
- Outcomes for neonates with confirmed SARS-CoV-2 infection are good; only 6.5% (6/92) of admitted neonates required intensive care, and COVID-19 was not the primary diagnosis recorded for these babies. There were no neonatal deaths among babies with confirmed infection.

## Introduction

Confirmed neonatal infection with Severe Acute Respiratory Syndrome Coronavirus 2 (SARS-CoV-2), defined as a positive viral test in the first 27 days after birth, is uncommon (1-4). A study in the UK identified 66 neonates with confirmed infection admitted to hospital between 1 March and 30 April 2020, giving an estimated infection/admission rate of 5.6/10,000 live births (1). Less than 2% of babies born to women with confirmed infection around the time of birth go on to develop confirmed infection themselves (5). However, babies born to women with confirmed infection are more likely to be born prematurely or be admitted to the neonatal unit, regardless of infant SARS-CoV-2 status (5-8). Neonates with confirmed infection can develop severe disease, however reports on the proportion of SARS-CoV-2 positive neonates requiring admission to intensive care vary depending on the definition of intensive care (1, 2, 13, 14).

To date, most neonatal SARS-CoV-2 studies have focused on the risk and consequences of transmission to the neonate from an infected mother (5, 15). However, in the neonatal period, babies are exposed to multiple other potential sources of infection, for example other caregivers, healthcare professionals, and the general population. Previous studies have reported on the rates and outcomes of neonates admitted to hospital with a positive SARS-CoV-2 test (1), however, population level data including those testing positive in the community are lacking (4). The aim of this study was to examine all confirmed cases of SARS-CoV-2 infection in infants aged 0-27 days in Scotland from March 2020 until January 2022.

## Methods

### Study population

Data were obtained from the “COVID-19 in pregnancy in Scotland” (COPS) study dataset as updated in mid-May 2022 (16, 17). COPS contains data on all ongoing and completed pregnancies to women in Scotland, and live born babies resulting from those pregnancies, from 1 January 2015 onwards. The dataset can be linked to medical records including SARS-CoV-2 viral testing, admissions to neonatal and paediatric care, and deaths (16); further details can be found in Supplemental file 1. For this study, we included all live born babies born in Scotland between 1 March 2020 and 31 January 2022 who had a valid Community Health Index (CHI) number available (required for data linkage) within the COPS dataset.

### Identifying confirmed SARS-CoV-2 infections

COPS includes information on all SARS-CoV-2 viral tests undertaken on women and babies within the cohort, both Reverse Transcription Polymerase Chain Reaction (RT-PCR) SARS-CoV-2 tests and Lateral Flow Device (LFD) tests where the result has been logged on the UK Government website (16, 18). Up to and including 5 Jan 2022, confirmed SARS-CoV-2 infection was defined as a positive viral RT-PCR test result. From 6 Jan 2022 onwards, confirmed infection was defined as a positive viral RT-PCR or a positive LFD test (unless the positive LFD result was followed by a negative RT-PCR result within 48 hours). For any individual, the date that their first positive test sample was taken was used as the date of onset of their first episode of infection. Confirmed neonatal infection was defined as a positive test with date of onset from birth to 27 days old inclusive. Maternal infection at the time of birth was defined as a confirmed infection with date of onset in the 14 days leading to birth, on the day of birth, or the day after giving birth.

For all babies with confirmed neonatal infection, data were obtained from the COPS database regarding the age of the baby in days at date of onset of infection; maternal age, socioeconomic level, ethnicity, infection status at the time of birth; the baby’s sex and gestation at birth. Maternal socioeconomic level was based on the Scottish Index of Multiple Deprivation (SIMD) quintile (19).

### Identifying hospital admissions associated with confirmed neonatal SARS-CoV-2 infection

A hospital admission associated with confirmed neonatal SARS-CoV-2 infection was defined as an admission to neonatal or paediatric care where the date of onset of infection was in the 7 days prior to the date of admission or during the admission, or where COVID-19 was recorded as the main diagnosis (ICD10 code U07.1 or U07.2). Admissions to neonatal units were identified through the Scottish Birth Record (SBR, (20)), and admissions to paediatric wards through hospital inpatient and day-case discharge records (SMR01, (21)). SARS-CoV-2-associated admission records were analysed to identify the highest level of care provided in the neonatal unit (intensive care, high dependency, or special care) or whether the stay included an episode in a paediatric intensive care unit (PICU) (“Significant Facility” coded to 13, intensive care unit (22)); length of stay for the entire hospital stay; whether COVID-19 was listed as the main diagnosis; and whether the infection was likely to be nosocomial. A probable nosocomial infection was defined as when the first positive viral test was taken on day 7 or later of an ongoing admission.

### Calculation of rates and confidence intervals

Due to the relatively small numbers of cases, all data reported here are descriptive only with no formal statistical comparisons. Rates were calculated using the number of babies with confirmed neonatal infection and the total number of live births during the study time-period. The confidence intervals were calculated using Wilson score estimates. The analysis and generation of figures was carried out using R (3.6.1) and RStudio (1.1.463) and codes are available at https://github.com/Public-Health-Scotland/COPS-public.git.

### Ethics and reporting

COPS has ethical approval from the National Research Ethics Service Committee, South East Scotland 02 (REC 12/SS/0201: SA 2) and information governance approval from the Public Benefit and Privacy Panel for Health and Social Care (2021-0116). A pre-analysis study protocol was developed and made publicly available at https://github.com/Public-Health-Scotland/COPS-public.git. RECORD (23) and STROBE (24) were used to guide transparent reporting.

## Results

Overall, 92,032 live births in Scotland between 1 March 2020 and 31 January 2022 were included in the COPS dataset, of whom 92,009 had a valid CHI number. One-hundred and forty-two neonates with confirmed SARS-CoV-2 were identified from the national viral testing data. Of these, 141 neonates were within the COPS cohort and were included in the analysis. The remaining baby was presumed to have been born outside of Scotland and was excluded.

### Neonatal infection rates

Across the study period, the overall neonatal confirmed SARS-CoV-2 infection rate was 153 per 100,000 live births, however this varied by month from 0-665 per 100,000 live births (figure 1A, table S1). For context, Figure 1B shows the neonatal confirmed infection rate alongside the rates for older children (drawing on other population-based data held by Public Health Scotland) (Table S2). The neonatal infection rate was consistently the lowest, though all paediatric age groups showed similar peaks of infection in autumn 2021 and December 2021/January 2022. For additional context, the monthly rates of confirmed infection in pregnant women over the study period are presented in Table S3.

**Figure 1:**
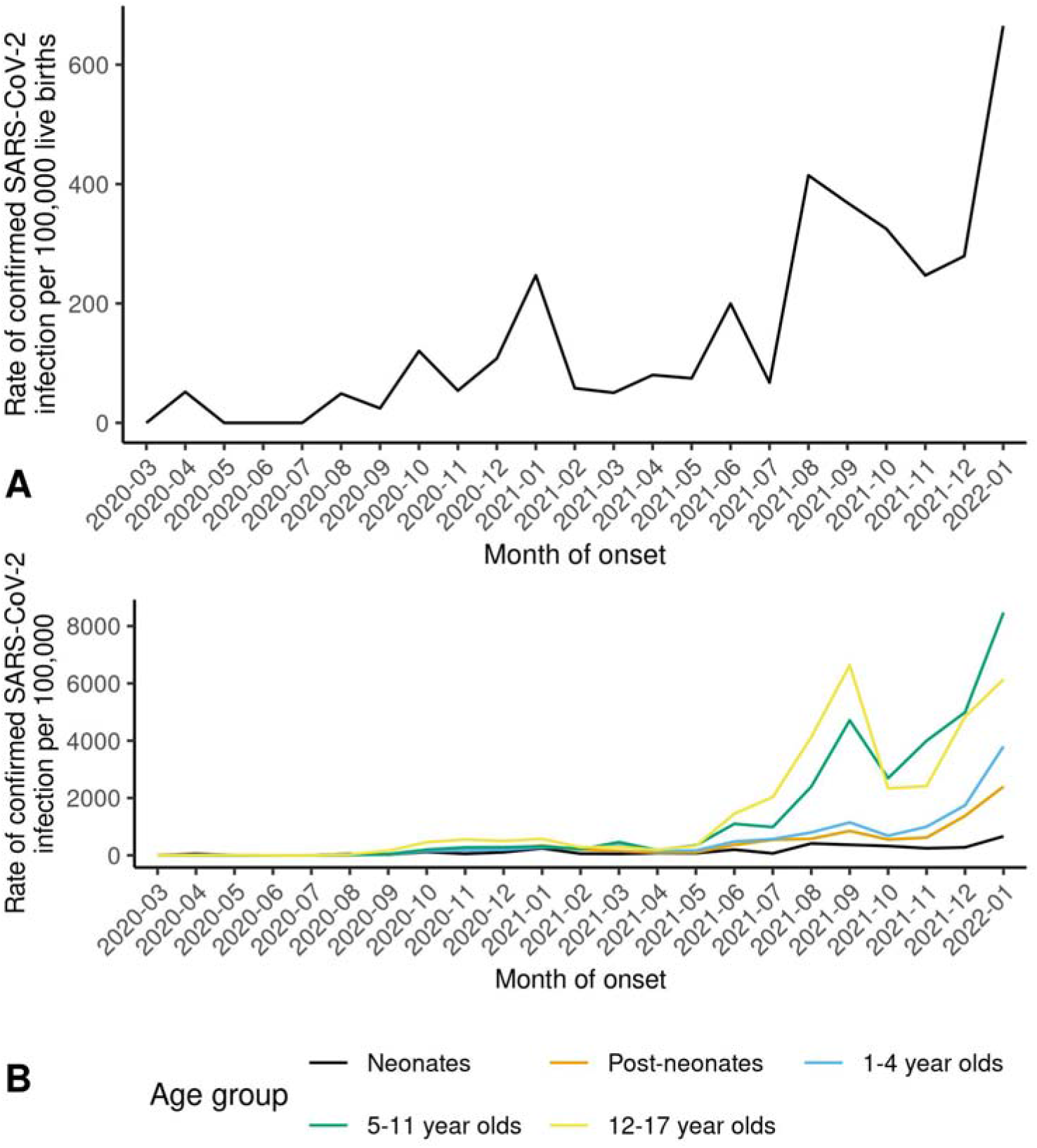
Scotland, March 2020 to January 2022. A Monthly rate of confirmed SARS-CoV-2 infection in neonates (babies aged 0-27 days) per 100,000 live births. B Monthly rates of confirmed SARS-CoV-2 infection in neonates, post neonates (babies aged 28-364 days), and children aged 1-4 years, 5-11 years and 12-17 years per 100,000 live births (neonates) or population (older age groups).

For further details on data sources and estimates see Table S2.

### Infant and maternal characteristics and confirmed neonatal SARS-CoV-2 infection

The infant and maternal characteristics of neonates with confirmed SARS-CoV-2 infection are shown in Tables 1 and 2 respectively. Rates of confirmed neonatal infection were highest among babies born to younger women and to women from more deprived areas, although confidence intervals overlapped. Rates of neonatal infection among babies born to women from minority ethnic groups were very uncertain due to low numbers. The rate of confirmed neonatal infection was substantially higher in babies born to women with (compared to without) confirmed infection at the time of birth, however the absolute risk of neonatal infection was low in both groups, and the majority of neonates with confirmed infection were born to mothers who did not have confirmed infection at birth (Table 2).

**Table 1:** Infant characteristics and confirmed neonatal SARS-CoV-2 infection.

|  | **Total live births** | **No. of neonates SARS-CoV-2 positive** | **Rate per 100,000 live births** | **Lower CI** | **Upper CI** |
| --- | --- | --- | --- | --- | --- |
| **Infant sex** |  |  |  |  |  |
| Male | 47,232 | 79 | 167.3 | 133.3 | 209.6 |
| Female | 44,777 | 62 | 138.5 | 107.1 | 178.7 |
| **Gestation at birth** |  |  |  |  |  |
| Preterm (22-36wk) | 7,231 | 17 | 235.1 | 141.5 | 384.7 |
| *Earlier preterm (22-33wk)* | *1,946* | *3* | *154.2* | *39.8* | *490.2* |
| *Later preterm (34-36wk)* | *5,285* | *14* | *264.9* | *150.8* | *456.0* |
| Term+ (37-44wk) | 84,735 | 124 | 146.3 | 122.2 | 175.1 |
| Unknown | 43 | 0 | - | - | - |
| **Total** | 92,009 | 141 | 153.3 | 129.5 | 181.3 |

CI = 95% confidence intervals. Wk = weeks.

**Table 2:** Maternal characteristics and confirmed neonatal SARS-CoV-2 infection.

|  | Total no.<br>of live<br>births | No. of<br>neonates<br>SARS-CoV-2<br>positive | Rate per<br>100,000 live<br>births | Lower CI | Upper CI |
| --- | --- | --- | --- | --- | --- |
| **Maternal age (years)** |  |  |  |  |  |
| ≤19 | 3,248 | 12 | 369.5 | 200.3 | 664.2 |
| 20-24 | 12,886 | 26 | 201.8 | 134.6 | 300.1 |
| 25-29 | 26,723 | 41 | 153.4 | 111.5 | 210.2 |
| 30-34 | 30,837 | 47 | 152.4 | 113.3 | 204.5 |
| 35-39 | 15,471 | 12 | 77.6 | 42.0 | 139.6 |
| ≥40 | 2,744 | 3 | 109.3 | 28.2 | 347.8 |
| Unknown | 100 | 0 | - | - | - |
| **Maternal deprivation level (SIMD quintile)** |  |  |  |  |  |
| 1 - most deprived | 21,189 | 47 | 221.8 | 164.8 | 297.5 |
| 2 | 18,501 | 29 | 156.8 | 107.0 | 228.2 |
| 3 | 16,860 | 21 | 124.6 | 79.1 | 193.9 |
| 4 | 19,374 | 26 | 134.2 | 89.5 | 199.6 |
| 5 - least deprived | 16,018 | 18 | 112.4 | 68.7 | 181.4 |
| Unknown | 67 | 0 | - | - | - |
| **Maternal ethnicity** |  |  |  |  |  |
| White | 77,481 | 118 | 152.3 | 126.6 | 183.0 |
| South Asian | 3,073 | 8 | 260.3 | 121.1 | 534.4 |
| Black/Caribbean/African | 1,431 | 3 | 209.6 | 54.2 | 666.0 |
| Mixed or other ethnic group | 3,222 | 5 | 155.2 | 57.1 | 384.4 |
| Unknown | 6,802 | 7 | 102.9 | 45.1 | 222.2 |
| **Maternal SARS-CoV-2 infection status at birth** |  |  |  |  |  |
| SARS-CoV-2 positive | 828 | 15 | 1811.6 | 1055.2 | 3041.7 |
| Not SARS-CoV-2 positive | 91,181 | 126 | 138.2 | 115.6 | 165.1 |
| **Total** | 92,009 | 141 | 153.3 | 129.5 | 181.3 |

CI = 95% confidence intervals. SIMD = Scottish Index of Multiple Deprivation

### Age in days at date of first positive test

The incidence of confirmed infection over the neonatal period followed a linear trend (figure 2, table S4). Of the 15 infants born to a woman with confirmed infection at birth, none tested positive at <2 days of age, 9 first tested positive between day 2 and 7, and 6 on day 8 or later.

**Figure 2:**
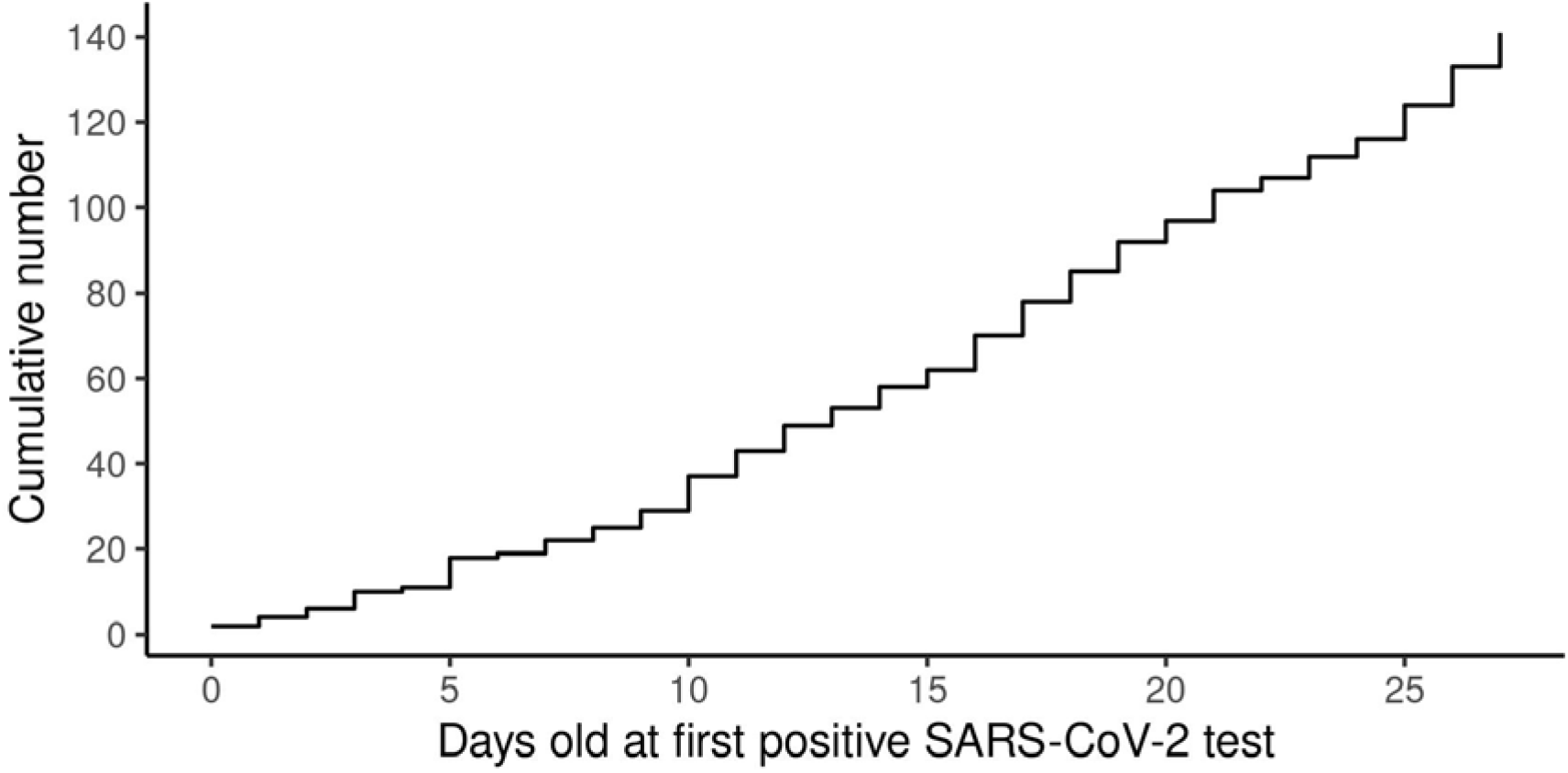
Cumulative number of confirmed SARS-CoV-2 infections in neonates by age at time of first positive test

### Hospital admission and outcomes of babies with confirmed neonatal SARS-CoV-2 infection

Of the 141 babies with confirmed neonatal infection, 92 (92/141, 65.2%) had at least one admission which was temporally associated with their positive SARS-CoV-2 test of whom 6 (6.5%) were admitted to neonatal and/or paediatric intensive care. Overall, 101 distinct SARS-CoV-2 associated admissions to neonatal and/or paediatric care were identified. None of the six SARS-CoV-2-associated admissions to a neonatal unit had COVID-19 coded as the main diagnosis underlying the admission, and three involved probable nosocomial infection. By contrast, 66% (64/97) of the SARS-CoV-2-associated admissions to paediatric care had COVID-19 coded as the main diagnosis, and only one involved probable nosocomial infection. None of the six admissions to paediatric intensive care had COVID-19 coded as the main diagnosis (Table 3).

**Table 3:** Hospital admissions temporally associated with a positive SARS-CoV-2 test among babies with confirmed neonatal infection.

| Admissions to neonatal unit (N=6) | COVID-19 coded as main diagnosis |  | All |
| --- | --- | --- | --- |
|  | Yes | No |  |
| Maximum level of care |  |  |  |
| Intensive care | 0 | 2 | 2 |
| High dependency or special care only | 0 | 4 | 4 |
| Probable nosocomial infection |  |  |  |
| Yes | 0 | 3 | 3 |
| No | 0 | 3 | 3 |
| Total | 0 | 6 | 6 |
| Mean LOS (median, IQR) | NA | 28.8,(22.5, 17.8) | 28.8, (22.5, 17.8) |
| Admissions/transfers to paediatric care (N= 97) | COVID-19 coded as main diagnosis |  | All |
|  | Yes | No |  |
| Maximum level of care |  |  |  |
| PICU | 0 | 6 | 6 |
| No PICU | 64 | 27 | 91 |
| Probable nosocomial infection |  |  |  |
| Yes | 0 | 1 | 1 |
| No | 64 | 32 | 96 |
| Total | 64 | 33 | 97 |
| Mean LOS (median, IQR) | 1.6, (1, 2) | 4.1, (2, 2) | 2.4, (1, 2) |

N=101 separate admissions to neonatal or paediatric care of 92 neonates (2 admissions involved a transfer from neonatal to paediatric care).
PICU = paediatric intensive care unit, LOS = length of stay, IQR = interquartile range, NA = not applicable.

Over the two-year study period, the proportion of babies with confirmed neonatal infection that were admitted to hospital remained broadly consistent (Table S5 and Figure S1).

There were no neonatal deaths among the 141 babies with confirmed neonatal infection. This compares to a background neonatal mortality rate in March 2020 to January 2022 of 2.2/1,000 live births (206/91,864, 95% CI 2.0-2.6) among uninfected babies.

## Discussion

These results show that confirmed neonatal SARS-CoV-2 infection is uncommon, with only 141 babies in Scotland aged 27 days or under having confirmed infection between March 2020 and January 2022. Broadly speaking, the secular trend in the neonatal infection rate followed that seen in older age groups, albeit at much lower levels, and factors associated with higher infection rates among pregnant women, such as young maternal age and living in a more deprived area, were also associated with higher neonatal infection rates. The rate of neonatal SARS-CoV-2 infection in babies born to women with confirmed infection at the time of birth was over ten times that of babies of women without confirmed infection, however even in these babies, SARS-CoV-2 infection remained uncommon. Two-thirds of neonates with confirmed SARS-CoV-2 infection were admitted to hospital, primarily to paediatric care. However only 6.5% of admitted babies required (paediatric or neonatal) intensive care and none of these babies had COVID-19 coded as their main diagnosis.

These data align with previous studies on admission rates for neonatal SARS-CoV-2 (1, 4), and give a useful insight into the total burden of confirmed neonatal infection in the UK both in hospital and the community. A key strength of this study is that it takes a population level view of neonatal SARS-CoV-2 infection, rather than confining results to only those born to infected women, or only those admitted to hospital. Our data also encompasses almost two years of the pandemic, including the emergence of new variants such as Omicron and the widespread availability of COVID-19 vaccines. In keeping with the published rates of mother to infant transmission (5), we found just under 2% of babies born to women positive for SARS-CoV-2 at birth also had a positive test at some point in the neonatal period. Not all babies of infected women were tested at birth (see below), and it is possible that rates may have been higher if all were screened (10).

A significant limitation is that this was not a screening study, therefore we cannot claim to have captured every neonate with SARS-CoV-2. Testing procedures for neonates born to an infected woman and/or admitted to a neonatal unit varied both nationally and internationally (26) and changed significantly over the study period. Access to community-based testing was limited early in the pandemic, with RT-PCR testing becoming widely available (including for children) from August 2020 and free home LFD testing from April 2021. Testing policy for inpatients has also evolved and varies over time (and by unit) for both neonatal units and for paediatric admissions. These factors are likely to have resulted in differences in case ascertainment over time, with some cases being missed, particularly in the early part of the pandemic. During the study period, infants may have been tested for several reasons including having clinical signs of SARS-CoV-2 infection, having close contact with a case, or on admission to hospital (27). However, infants who did not develop clinical signs may have been missed and it is likely that not every baby with clinical signs was tested in the community. Around half of neonates may appear well or show mild clinical signs which may not have prompted testing (1, 2, 9-13). It is therefore likely that we have underestimated the actual number of neonates with SARS-CoV-2.

Reassuringly, we demonstrate that the clinical outcomes of neonates with confirmed SARS-CoV-2 infection are good, with no deaths and no intensive care admissions for which COVID-19 was the main diagnosis. Other studies have recorded higher rates of critical care admissions; Swann et al reported that up to 33% of neonates with confirmed infection in the first wave of SARS-CoV-2 in the UK required critical care (14), and a subsequent study over a longer time period found that 20% of neonates required critical care (28). However, these studies only included admitted babies (with those cared for at home excluded), and the definition of critical care used included admission to a PICU or any level of care in a neonatal unit (28 and personal communication). Our data suggest that a much lower proportion of neonates with SARS-CoV-2 infection truly require intensive care.

Despite these positive outcomes, a very high proportion 92 (92/141, 65.2%) of neonates with confirmed SARS-CoV-2 infection had a hospital admission around the time of their positive viral test. This is perhaps not surprising, as fever is a common clinical sign of SARS-CoV-2 infection in neonates (1, 2, 9-13) and according to UK guidelines (30), a temperature above 38°C should prompt blood and urine tests, lumbar puncture, and administration of intravenous antibiotics. This demonstrates the indirect effects of SARS-CoV-2 on infants who may receive additional invasive investigations and treatments aimed at potential bacterial infections, although their clinical signs have a viral cause. This study was limited in that detailed information on signs and treatments received in hospital were lacking. However, detailed information on the care of babies admitted with SARS-CoV-2 early in the pandemic is available through the British Paediatric Surveillance Unit (31).

In summary, confirmed SARS-CoV-2 infection in the neonatal period has been uncommon throughout the pandemic in Scotland to date, even among babies born to women with confirmed infection at the time of birth. A high proportion of neonates with confirmed infection are admitted, and though outcomes are generally good, the potential effect of this on the baby, family, and services may be considerable. Continued data collection and analysis will be important to assess the ongoing impact of SARS-CoV-2 in the neonatal population as testing and isolation requirements continue to evolve, and new viral variants potentially emerge.

## Supporting information

STROBE

## Data Availability

Patient-level data underlying this article cannot be shared publicly due to data protection and confidentiality requirements. Data access for approved researchers can be authorised by the Public Benefit and Privacy Panel for Health and Social Care (https://www.informationgovernance.scot.nhs.uk/pbpphsc/). Enquiries regarding data availability should be directed to.

https://github.com/Public-Health-Scotland/COPS-public.git

## STROBE checklist

Provided as a supplemental file.

## Contributor statement

Conceptualization, RW; Data curation, AG, LL; Formal Analysis, AG, LL, FM ; Methodology, RW, AG, LL; Interpretation, AG, JK, LP, FM, LF, CG; Project administration, AG, FM ; Visualization, AG; Supervision, RW; Writing – Original Draft, FM; Writing –Review & Editing, FM, AG, LL, RW, CS, JK, LP, UA, BA, CC, JC, CD, JD, SH, LH, LeH, TM, EM, BT, LF, CG, CM, LR, AS, SS. AG and FM are joint first authors.

## Licence statement

I, the Submitting Author has the right to grant and does grant on behalf of all authors of the Work (as defined in the below author licence), an exclusive licence and/or a non-exclusive licence for contributions from authors who are: i) UK Crown employees; ii) where BMJ has agreed a CC-BY licence shall apply, and/or iii) in accordance with the terms applicable for US Federal Government officers or employees acting as part of their official duties; on a worldwide, perpetual, irrevocable, royalty-free basis to BMJ Publishing Group Ltd (“BMJ”) its licensees and where the relevant Journal is co-owned by BMJ to the co-owners of the Journal, to publish the Work in Archives of Disease in Childhood and any other BMJ products and to exploit all rights, as set out in our licence.

The Submitting Author accepts and understands that any supply made under these terms is made by BMJ to the Submitting Author unless you are acting as an employee on behalf of your employer or a postgraduate student of an affiliated institution which is paying any applicable article publishing charge (“APC”) for Open Access articles. Where the Submitting Author wishes to make the Work available on an Open Access basis (and intends to pay the relevant APC), the terms of reuse of such Open Access shall be governed by a Creative Commons licence – details of these licences and which Creative Commons licence will apply to this Work are set out in our licence referred to above.

