## Supplemental for "Confirmed SARS-CoV-2 infection in Scottish neonates 2020-2022: a national, population-based cohort study"

**Supplementary data and figures**

**Supplementary methods**

**Study population**

We derived and analysed data came from the “COVID-19 in pregnancy in Scotland” (COPS) study dataset as updated in mid-May 2022. The COPS dataset is described in detail elsewhere (1, 2). In brief, it comprises a population-based, dynamic cohort which includes data on all ongoing and completed pregnancies to women in Scotland, and live born babies resulting from those pregnancies, from 1 January 2015 onwards. The dataset is based on linkage of health service and statutory national datasets, including those relating to pregnancy-related care and births, SARS-CoV-2 viral testing and COVID-19 vaccinations, admissions to neonatal and paediatric care, and deaths (1). All individuals in Scotland who receive care from the National Health Service (NHS) (which will include essentially all pregnant women and neonates as there is no private maternity care in Scotland) are assigned a unique patient identifier, the Community Health Index (CHI) number (3) which allows for linkage of medical records relating to an individual. Within the COPS database, the CHI number for both the mother and the baby are included on NHS live birth notification records, further allowing intergenerational linkage of records relating to mothers and their babies. For this study, we included all live born babies born in Scotland between 1^st^ March 2020 and 31^st^ January 2022 who had a valid CHI number available within the COPS dataset.

**Identifying confirmed SARS-CoV-2 infections**

COPS includes information on all SARS-CoV-2 viral tests undertaken on women and babies within the cohort from the NHS Scotland Corporate Data Warehouse. The warehouse includes results of all Reverse Transcription Polymerase Chain Reaction (RT-PCR) SARS-CoV-2 tests processed through NHS Scotland and UK Government Regional Testing Centre ('Lighthouse') laboratories. It also includes information on all SARS-CoV-2 Lateral Flow Device (LFD) tests where the result has been logged by the individual taking the test (or their parent/carer) on the UK Government website (1, 4). Up to and including 6 Jan 2022, confirmed SARS-CoV-2 infection was defined as a positive viral RT-PCR test result.  From 6 Jan 2022 onwards, confirmed infection was defined as a positive viral RT-PCR test result or a positive LFD test result (unless the positive LFD result was followed by a negative viral RT-PCR result within 48 hours). This is consistent with the contemporaneous case definition in Scotland (5).

For any individual, the date that their first positive test sample was taken was used as the date of onset of their first episode of infection. Information on the presence of clinical signs, and date of onset of signs, was not always available on testing records. Subsequent positive samples taken within 90 days of a first positive sample were discounted.  A subsequent positive sample taken more than 90 days after a first positive result was taken as indicating a subsequent confirmed infection.

Confirmed neonatal infection was defined as a confirmed infection with date of onset at any point from birth to 27 days old inclusive. By this definition, if infected each baby can therefore only have one episode of confirmed infection during the neonatal period.  Maternal infection at the time of childbirth was defined as a confirmed infection with date of onset in the 14 days leading to birth, on the day of childbirth, or on the day after giving birth.

For all babies with confirmed neonatal infection, data were obtained from the COPS database regarding: the age of the baby in days at date of onset of infection; maternal age, socioeconomic status, ethnicity, infection status at the time of childbirth; the baby’s sex and gestation at birth. Maternal socioeconomic status was based on the Scottish index of multiple deprivation (SIMD) quintile. SIMD is an area-based measure of material deprivation derived from the postcode of residence (6)).

**Identifying admissions of neonates with SARS-CoV-2**

Neonates requiring hospital inpatient care may be cared for alongside their mother in a postnatal ward or admitted to a neonatal unit or paediatric ward (including paediatric intensive care units, PICU). Babies who have been discharged home after birth - who then require readmission to hospital -will generally be admitted to a paediatric ward (rather than a neonatal unit) to avoid importing and transmitting infections to neonatal units.

We first identified all admissions to neonatal units and paediatric wards in the neonatal period (date of admission from birth to 27 days old inclusive) for babies in our study population. Admissions to neonatal units were identified through the Scottish Birth Record (SBR, (7)), and admissions to paediatric wards through the Scottish Morbidity Record hospital inpatient and daycase discharge records (SMR01, (8)). A stay in neonatal care was defined as any SBR admission record which included at least one episode in intensive care, high dependency care, or special care. Any SMR01 record in the relevant period was included as a paediatric admission. Of note, this method does not capture neonates who were readmitted from home to a postnatal unit, rather than to a neonatal unit or paediatric care. Therefore, it is possible that some admissions may have been missed.

For babies with confirmed neonatal SARS-CoV-2 infection, we then identified SARS-CoV-2-associated admissions for further analysis. A SARS-CoV-2 associated admission was defined as an admission to neonatal or paediatric care where the date of onset of infection was in the 7 days prior to the date of admission or during the admission, or where COVID-19 was recorded as the main diagnosis (ICD10 code U07.1 or U07.2). For this more detailed analysis, any admission in the 7-day period following an infection in the neonatal period was included in the analysis even if the admission itself occurred after the neonatal period.

Neonatal and paediatric admission records for these SARS-CoV-2-associated admissions were analysed to identify the highest level of care provided in the neonatal unit (intensive care, high dependency or special care) or whether the stay included an episode in a paediatric intensive care unit, length of stay for the entire hospital stay, whether COVID-19 was listed as the main diagnosis, and whether the infection was likely to be nosocomial. A probable nosocomial infection is defined as when the first positive test was taken on day 7 of an ongoing admission or later.

**Calculation of rates and confidence intervals**

Due to the relatively small numbers involved, all data reported here are descriptive only and no formal statistical comparisons have been made. Rates were calculated using the number of babies with confirmed neonatal infection (numerator) and the total number of live births (denominator) during the relevant time-period (individual months or the full study period 1 March 2020 to 31 January 2022), with confidence intervals calculated using Wilson score estimates. This approach to calculating rates supports production of the timeliest results, however it does allow a mismatch between numerator and denominator. For example, in any one month some of the babies with confirmed infection during that month may have been born in the previous month. Given the number of live births is fairly consistent month to month, we believe that this mismatch should have minimal impact on the interpretation of our findings.

5.	Public Health Scotland PH. COVID-19 - guidance for health protection teams (HPTs). 2022.

6.	Scottish Government S. Introducing the Scottish Index of Multiple Deprivation 2020. National statistics publication; 2020.

7.	Information Services Division Scotland. Scottish Birth Record : ISD Scotland Data

Dictionary	. 2022 [Available from: <https://www.ndc.scot.nhs.uk/Data-Dictionary/SMR-Datasets/Scottish-Birth-Record/>.

8.	Information Services Division Scotland. SMR01: ISD Data dictionary 2022 [Available from: <https://www.ndc.scot.nhs.uk/Dictionary-A-Z/Definitions/index.asp?Search=S&ID=460&Title=SMR01%20-%20General/Acute%20Inpatient%20and%20Day%20Case>.

9.	National Records of Scotland. Mid-2020 Population Estimates Scotland. 2021.

**Table S1**

Monthly rate of confirmed SARS-CoV-2 infection in babies aged 27 days and under per 100,000 live births. CI = 95% confidence interval.

| **Month** | **Total number of live births** | **Number of confirmed SARS-CoV-2 infections in neonates with date of onset in month** | **Overall rate of confirmed SARS-CoV-2 infection in neonates**  **(per 100,000 live births)** | **Lower CI** | **Upper CI** |
| --- | --- | --- | --- | --- | --- |
| **Mar-20** | 4,001 | 0 | 0.0 | 0.0 | 119.6 |
| **Apr-20** | 3,857 | 2 | 51.9 | 9.0 | 208.9 |
| **May-20** | 3,888 | 0 | 0.0 | 0.0 | 123.1 |
| **Jun-20** | 4,083 | 0 | 0.0 | 0.0 | 117.2 |
| **Jul-20** | 4,282 | 0 | 0.0 | 0.0 | 111.8 |
| **Aug-20** | 4,078 | 2 | 49.0 | 8.5 | 197.6 |
| **Sep-20** | 4,106 | 1 | 24.4 | 1.3 | 158.0 |
| **Oct-20** | 4,152 | 5 | 120.4 | 44.3 | 298.4 |
| **Nov-20** | 3,703 | 2 | 54.0 | 9.4 | 217.6 |
| **Dec-20** | 3,707 | 4 | 107.9 | 34.6 | 296.4 |
| **Jan-21** | 3,641 | 9 | 247.2 | 120.7 | 487.1 |
| **Feb-21** | 3,447 | 2 | 58.0 | 10.1 | 233.7 |
| **Mar-21** | 3,966 | 2 | 50.4 | 8.7 | 203.2 |
| **Apr-21** | 3,742 | 3 | 80.2 | 20.7 | 255.2 |
| **May-21** | 4,022 | 3 | 74.6 | 19.3 | 237.4 |
| **Jun-21** | 4,006 | 8 | 199.7 | 92.9 | 410.1 |
| **Jul-21** | 4,445 | 3 | 67.5 | 17.4 | 214.9 |
| **Aug-21** | 4,342 | 18 | 414.6 | 253.5 | 668.5 |
| **Sep-21** | 4,339 | 16 | 368.8 | 218.3 | 612.4 |
| **Oct-21** | 4,308 | 14 | 325.0 | 185.0 | 559.3 |
| **Nov-21** | 4,048 | 10 | 247.0 | 125.6 | 470.1 |
| **Dec-21** | 3,937 | 11 | 279.4 | 147.0 | 515.9 |
| **Jan-22** | 3,909 | 26 | 665.1 | 443.9 | 988.0 |
| **Total** | 92,009 | 141 | 153.3 | 129.5 | 181.3 |

**Table S2**

Rates of confirmed SARS-CoV-2 infection in post neonatal infants and children up to 17 years of age. Estimated number of children in each age group (denominator for rates) taken from National Records of Scotland, Mid-2020 Population Estimates Scotland and is kept constant throughout (9). Number of post neonatal infants (under 1 year) estimated using the Mid-2020 Population Estimate minus 4,000 (the average number of live births per month). Number of positive tests obtained from ECOSS (4).

| **Month** | **Estimated no. of post-neonates, under 1yr**  **(d28 to d364)** | **No. confirmed SARS-CoV-2 infection post-neonates** | **Overall rate post-neonates (per 100,000)** | **Estimated no. of children aged 1-4 years** | **No. confirmed SARS-CoV-2 infection aged 1-4 years** | **Overall rate aged 1-4 years (per 100,000)** | **Estimated no. of children aged 5-11 years** | **No. confirmed SARS-CoV-2 infection aged 5-11 years** | **Overall rate children aged 5-11 years (per 100,000)** | **Estimated no. of children aged 12-17 years** | **No. confirmed SARS-CoV-2 infection children aged 12-17 years** | **Overall rate of children aged 12-17 years (per 100,000)** |
| --- | --- | --- | --- | --- | --- | --- | --- | --- | --- | --- | --- | --- |
| **Mar-20** | 44,646 | 8 | 17.9 | 215,171 | 8 | 3.7 | 418,842 | 8 | 1.9 | 344,274 | 12 | 3.5 |
| **Apr-20** | 44,646 | 1 | 2.2 | 215,171 | 14 | 6.5 | 418,842 | 23 | 5.5 | 344,274 | 39 | 11.3 |
| **May-20** | 44,646 | 1 | 2.2 | 215,171 | 4 | 1.9 | 418,842 | 27 | 6.6 | 344,274 | 44 | 12.8 |
| **Jun-20** | 44,646 | 0 | 0.0 | 215,171 | 3 | 1.4 | 418,842 | 5 | 1.2 | 344,274 | 11 | 3.2 |
| **Jul-20** | 44,646 | 0 | 0.0 | 215,171 | 2 | 0.9 | 418,842 | 7 | 1.7 | 344,274 | 2 | 0.6 |
| **Aug-20** | 44,646 | 10 | 22.4 | 215,171 | 27 | 12.6 | 418,842 | 55 | 13.1 | 344,274 | 115 | 33.4 |
| **Sep-20** | 44,646 | 11 | 24.6 | 215,171 | 67 | 31.1 | 418,842 | 178 | 42.5 | 344,274 | 550 | 159.8 |
| **Oct-20** | 44,646 | 71 | 159.0 | 215,171 | 323 | 150.1 | 418,842 | 828 | 197.7 | 344,274 | 1,580 | 458.9 |
| **Nov-20** | 44,646 | 77 | 172.5 | 215,171 | 363 | 168.7 | 418,842 | 1,164 | 277.9 | 344,274 | 1,903 | 552.8 |
| **Dec-20** | 44,646 | 101 | 226.2 | 215,171 | 420 | 195.2 | 418,842 | 1,176 | 280.8 | 344,274 | 1,733 | 503.4 |
| **Jan-21** | 44,646 | 154 | 344.9 | 215,171 | 599 | 278.4 | 418,842 | 1,322 | 315.6 | 344,274 | 1,977 | 574.3 |
| **Feb-21** | 44,646 | 89 | 199.4 | 215,171 | 524 | 243.5 | 418,842 | 892 | 213.0 | 344,274 | 970 | 281.8 |
| **Mar-21** | 44,646 | 67 | 150.1 | 215,171 | 768 | 356.9 | 418,842 | 1,932 | 461.3 | 344,274 | 1,038 | 301.5 |
| **Apr-21** | 44,646 | 41 | 91.8 | 215,171 | 296 | 137.6 | 418,842 | 755 | 180.3 | 344,274 | 645 | 187.4 |
| **May-21** | 44,646 | 44 | 98.6 | 215,171 | 363 | 168.7 | 418,842 | 1,465 | 349.8 | 344,274 | 1,135 | 329.7 |
| **Jun-21** | 44,646 | 162 | 362.9 | 215,171 | 1,039 | 482.9 | 418,842 | 4,602 | 1098.7 | 344,274 | 5,003 | 1453.2 |
| **Jul-21** | 44,646 | 242 | 542.0 | 215,171 | 1,225 | 569.3 | 418,842 | 4,128 | 985.6 | 344,274 | 7,011 | 2036.5 |
| **Aug-21** | 44,646 | 258 | 577.9 | 215,171 | 1,720 | 799.4 | 418,842 | 10,034 | 2395.7 | 344,274 | 14,200 | 4124.6 |
| **Sep-21** | 44,646 | 380 | 851.1 | 215,171 | 2,470 | 1147.9 | 418,842 | 19,740 | 4713.0 | 344,274 | 22,843 | 6635.1 |
| **Oct-21** | 44,646 | 247 | 553.2 | 215,171 | 1,470 | 683.2 | 418,842 | 11,279 | 2692.9 | 344,274 | 8,054 | 2339.4 |
| **Nov-21** | 44,646 | 277 | 620.4 | 215,171 | 2,149 | 998.7 | 418,842 | 16,748 | 3998.6 | 344,274 | 8,311 | 2414.1 |
| **Dec-21** | 44,646 | 615 | 1377.5 | 215,171 | 3,753 | 1744.2 | 418,842 | 20,901 | 4990.2 | 344,274 | 16,654 | 4837.4 |
| **Jan-22** | 44,646 | 1,071 | 2398.9 | 215,171 | 8,182 | 3802.6 | 418,842 | 35,480 | 8471.0 | 344,274 | 21,149 | 6143.1 |
| **Total** | 44,646 | 3,927 | 8795.9 | 215,171 | 25,789 | 11985.4 | 418,842 | 132,749 | 31694.3 | 344,274 | 114,979 | 33397.5 |

**Table S3:**

Rates of confirmed SARS-CoV-2 infection in pregnancy over study period. Data obtained from COPS cohort.

| **Month** | **Estimated No. women in Scotland with an ongoing pregnancy at start of month*** | **No. women with confirmed SARS-CoV-2 infection in pregnancy with date of onset in month**** | **Rate per 100,000 pregnant women***** |
| --- | --- | --- | --- |
| **Mar-20** | 38,018 | 18 | 47.4 |
| **Apr-20** | 37,557 | 35 | 93.2 |
| **May-20** | 37,017 | 23 | 62.1 |
| **Jun-20** | 36,698 | 2 | 5.5 |
| **Jul-20** | 36,536 | 3 | 8.2 |
| **Aug-20** | 36,183 | 14 | 38.7 |
| **Sep-20** | 36,086 | 61 | 169.0 |
| **Oct-20** | 36,003 | 286 | 794.4 |
| **Nov-20** | 36,254 | 261 | 719.9 |
| **Dec-20** | 36,837 | 324 | 879.6 |
| **Jan-21** | 37,647 | 476 | 1264.4 |
| **Feb-21** | 38,311 | 235 | 613.4 |
| **Mar-21** | 38,773 | 189 | 487.5 |
| **Apr-21** | 38,490 | 90 | 233.8 |
| **May-21** | 38,626 | 145 | 375.4 |
| **Jun-21** | 38,491 | 488 | 1267.8 |
| **Jul-21** | 38,363 | 796 | 2074.9 |
| **Aug-21** | 37,893 | 820 | 2164.0 |
| **Sep-21** | 37,229 | 975 | 2618.9 |
| **Oct-21** | 36,708 | 634 | 1727.1 |
| **Nov-21** | 36,131 | 688 | 1904.2 |
| **Dec-21** | 36,169 | 2,436 | 6735.1 |
| **Jan-22** | 36,514 | 2,986 | 8177.7 |
| **Total** | 151,230 | 10,552 | 6,977.5 |

* The total for this column is the number of women who were pregnant at some point during the study period.

** The total for this column is the number of women with confirmed SARS-CoV-2 infection in pregnancy during the study period.

*** The total for this column is the overall rate of confirmed SARS-CoV-2 infection in pregnancy during the study period.

**Table S4**

Date of onset of first positive test for neonates who had a confirmed SARS-CoV-2 infection at or below 27 days of age.

| **Date of onset of infection** | **Number of confirmed SARS-CoV-2 infection in neonates** | **Cumulative number of confirmed SARS-CoV-2 infection in neonates** |
| --- | --- | --- |
| d0 (date of birth) | 2 | 2 |
| d1 | 2 | 4 |
| d2 | 2 | 6 |
| d3 | 4 | 10 |
| d4 | 1 | 11 |
| d5 | 7 | 18 |
| d6 | 1 | 19 |
| d7 | 3 | 22 |
| d8 | 3 | 25 |
| d9 | 4 | 29 |
| d10 | 8 | 37 |
| d11 | 6 | 43 |
| d12 | 6 | 49 |
| d13 | 4 | 53 |
| d14 | 5 | 58 |
| d15 | 4 | 62 |
| d16 | 8 | 70 |
| d17 | 8 | 78 |
| d18 | 7 | 85 |
| d19 | 7 | 92 |
| d20 | 5 | 97 |
| d21 | 7 | 104 |
| d22 | 3 | 107 |
| d23 | 5 | 112 |
| d24 | 4 | 116 |
| d25 | 8 | 124 |
| d26 | 9 | 133 |
| d27 | 8 | 141 |

**Table S5**

Admission rates for neonates with confirmed SARS-CoV-2 infection over the study period. Rounded to 1 decimal place.

| **Month** | **Total number of live births** | **No. neonates with confirmed SARS-CoV-2 infection** | **No with a temporally associated admission** | **No with associated admission with COVID-19 as main diagnosis** | **Overall rate of confirmed SARS-CoV-2 infection in neonates (per 100,000 live births)** | **Rate of confirmed neonatal infection with temporally associated admission (per 100,000 live births)** | **Rate of confirmed neonatal infection with associated admission with COVID-19 as main diagnosis (per 100,000 live births)** |
| --- | --- | --- | --- | --- | --- | --- | --- |
| **Mar-20** | 4,001 | 0 | 0 | 0 | 0.0 | 0.0 | 0.0 |
| **Apr-20** | 3,857 | 2 | 2 | 0 | 51.9 | 51.9 | 0.0 |
| **May-20** | 3,888 | 0 | 0 | 0 | 0.0 | 0.0 | 0.0 |
| **Jun-20** | 4,083 | 0 | 0 | 0 | 0.0 | 0.0 | 0.0 |
| **Jul-20** | 4,282 | 0 | 0 | 0 | 0.0 | 0.0 | 0.0 |
| **Aug-20** | 4,078 | 2 | 0 | 0 | 49.0 | 0.0 | 0.0 |
| **Sep-20** | 4,106 | 1 | 1 | 1 | 24.4 | 24.4 | 24.4 |
| **Oct-20** | 4,152 | 5 | 1 | 1 | 120.4 | 24.1 | 24.1 |
| **Nov-20** | 3,703 | 2 | 2 | 2 | 54.0 | 54.0 | 54.0 |
| **Dec-20** | 3,707 | 4 | 1 | 1 | 107.9 | 27.0 | 27.0 |
| **Jan-21** | 3,641 | 9 | 3 | 3 | 247.2 | 82.4 | 82.4 |
| **Feb-21** | 3,447 | 2 | 0 | 0 | 58.0 | 0.0 | 0.0 |
| **Mar-21** | 3,966 | 2 | 2 | 0 | 50.4 | 50.4 | 0.0 |
| **Apr-21** | 3,742 | 3 | 1 | 0 | 80.2 | 26.7 | 0.0 |
| **May-21** | 4,022 | 3 | 2 | 2 | 74.6 | 49.7 | 49.7 |
| **Jun-21** | 4,006 | 8 | 6 | 4 | 199.7 | 149.8 | 99.9 |
| **Jul-21** | 4,445 | 3 | 3 | 2 | 67.5 | 67.5 | 45.0 |
| **Aug-21** | 4,342 | 18 | 13 | 8 | 414.6 | 299.4 | 184.3 |
| **Sep-21** | 4,339 | 16 | 13 | 11 | 368.8 | 299.6 | 253.5 |
| **Oct-21** | 4,308 | 14 | 7 | 5 | 325.0 | 162.5 | 116.1 |
| **Nov-21** | 4,048 | 10 | 8 | 6 | 247.0 | 197.6 | 148.2 |
| **Dec-21** | 3,937 | 11 | 6 | 2 | 279.4 | 152.4 | 50.8 |
| **Jan-22** | 3,909 | 26 | 21 | 12 | 665.1 | 537.2 | 307.0 |
| **Total** | 92,009 | 141 | 92 | 60 | 153.3 | 100.0 | 65.2 |

**Figure S1**

Trends in overall rate of confirmed neonatal SARS-CoV-2 infection compared to rate of neonatal infection with temporally associated admission, and with associated admission with COVID-19 coded as main diagnosis. Per 100,000 live births.

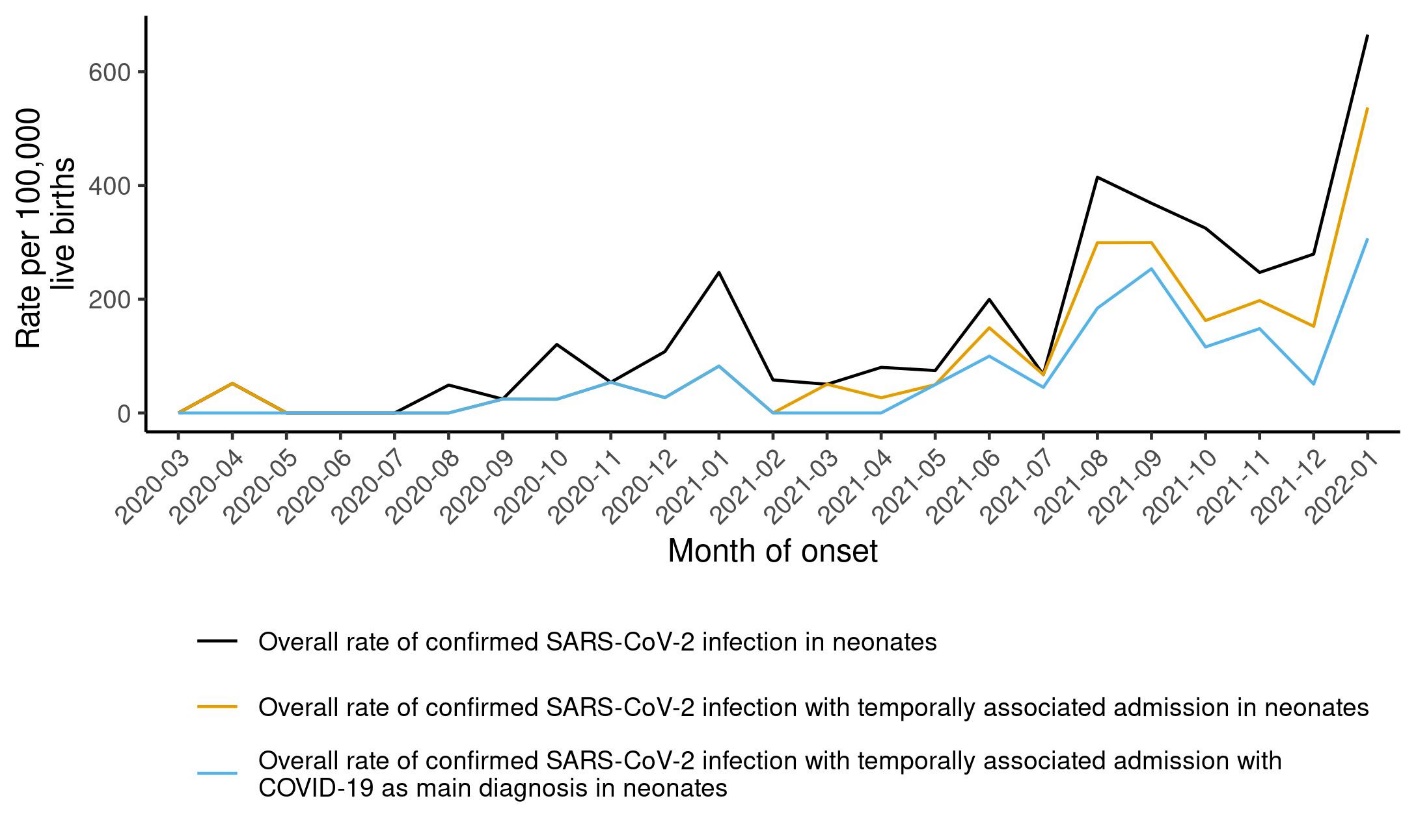
